# Association of opioid use disorder with healthcare utilization and cost in a public health system

**DOI:** 10.1101/2021.06.15.21258943

**Authors:** Oren Miron, Noam Barda, Ran Balicer, Ariel Kor, Shaul Lev-Ran

## Abstract

**Background and objectives:** To quantify the healthcare costs associated with opioid use disorder among members in a public healthcare system, and compare them to healthcare costs in the general population.

**Methods:** Retrospective cohort study in inpatient and outpatient care settings of Israel’s largest public healthcare provider (that covers 4.7 million members).

Participants included 1,173 members who had a diagnosis of opioid use disorder in the years between 2013 and 2018. Each patient was matched with 10 controls based on age and sex. The main outcome was monthly healthcare costs.

**Results:** The mean monthly healthcare cost of members with opioid use disorder was $1,102 compared to $211 among controls (5.2-fold difference; 95% confidence interval [CI]: 4.6-6.0). After excluding members with heroin related diagnoses before the index date (in order to focus on prescription opioids), this healthcare cost ratio did not substantially change (4.6-fold; 95%-CI: 3.9-5.4). Members with opioid use disorder under the age of 65 years had a cost difference of 6.1-fold (95%-CI: 5.2-7.1), while those 65 years and older experienced cost difference of 3.4-fold (95%-CI: 2.6-4.5). The category with the highest cost for members with opioid use disorder was inpatient services, which was 8.7-fold (95%-CI 7.2-10.4) greater than among controls.

**Conclusions:** Healthcare costs among individuals with opioid use disorder in a public health system were substantially higher than among controls, at least partially attributed to prescription opioid use disorder. Differences were greater among individuals younger than 65 years, highlighting the importance of preventing and treating opioid use disorder among younger adult populations.

## Introduction

In recent decades, the United States (US) has experienced a phenomenon of over-prescribing opioid medications, leading to a surge in both opioid use disorder and overdose.[1,2] Opioid use disorder is associated with health related harm, such as respiratory depression, digestive problems, and death.[3] Opioid use disorder is also related to other health hazards, including needle sharing for intravenous drug use and risky sexual practices, increasing the risk for hepatitis C and human immunodeficiency virus (HIV).[4]

The increase in opioid prescriptions in the US has also been associated with increased utilization of health care services. The annual cost of the population identified with opioid abuse in the US has been found to be 8 times higher than that of the general population, with 12 times more inpatient visits.[5–8]. Nevertheless, data pertaining to healthcare costs attributed to opioid use disorders in countries with a public healthcare system are lacking.

In recent years, an alarming increase in opioid prescriptions and the associated use disorder has occurred in other countries in the Organization for Economic Cooperation and Development (OECD). The largest opioid prescription increase in the OECD between 2011 and 2016 occurred in Israel, with a 125% increase; the second highest increase was in the United Kingdom. The Israeli increase in opioid consumption occurred mainly with Oxycodone and Fentanyl, which are highly addictive,[9] and are predominantly prescribed to non-elderly patients who do not suffer from malignancy, characteristics that have been found to increase the risk of developing opioid abuse.[10] There have been specific reports of increasing opioid use disorder in Israel, the United Kingdom, and other countries with public healthcare systems; yet, the health care burden of opioid abuse in these countries has been studied to a lesser extent than in the US.[11,12]

The current study examined differences in healthcare costs between individuals with opioid use disorders and matched controls between the years 2014 and 2018. The goal was to quantify the overall cost difference, as well as the cost difference among specific sub-groups, to determine the healthcare costs associated with opioid use disorder.

## Methods

### Study Setting and Population

The study was based on the data of Clalit Health Services (CHS), Israel’s largest integrated payer/provider healthcare organization, with 4.7 million patients (53% of the population). All clinical and administrative data on CHS patients is saved in an electronic health record database; prescribing and dispensing information is included, along with diagnoses and services received in both outpatient and inpatient settings.

### Study Design

This retrospective cohort study compared the healthcare costs of patients with opioid use disorder and their matched controls between January 1, 2014 and December 31, 2018. The study population included individuals 18 years and older who had continuous membership in CHS for least 12 months prior to the index date (to ensure sufficient data in the electronic medical record).

Patients with opioid use disorder were identified based on International Classification of Diseases, Ninth Revision (ICD-9) diagnosis codes (Opioid abuse-305.5; opioid dependence-304.0, 304.7) or diagnosis text denoting an opioid use disorder, as recorded in CHS’s inpatient and outpatient databases by the treating physician. The index date was set as the date of the first opioid use disorder diagnosis during the study period (January 1, 2014 to December 31, 2018).

Each patient with an opioid use disorder was individually matched to 10 other persons without an existing opioid use disorder diagnosis. Patients were matched based on age and sex. The index date for the control was set according to their matched patient’s index date.

The outcome of interest was healthcare costs as incurred by CHS. Costs were examined overall, and by categories of service, including inpatient care, emergency room care, prescription drug purchasing, outpatient care, laboratory testing, and imaging.

Costs were evaluated as a monthly rate. Each patient contributed time-under-observation 12 months following each opioid use diagnosis. Overlapping periods, which occurred if a patient was diagnosed several times within 12 months, led to a continuous time-under-observation. Patients who died during the follow-up period (within 12 months of a diagnosis) contributed time until their death. Controls contributed the same amount of time as their matched patients. Because patients diagnosed in the year preceding the study period could still contribute time-under-observation to the study, data were extracted between January 1, 2013 and December 31, 2018.

Data on each patient and their matched controls included sociodemographic information, clinical characteristics and comorbidities; sociodemographic and comorbidity data were collected current to the index date. Clinical characteristics were defined as the last values recorded prior to the index date.

### Statistical Analysis

Patients were described in regards to socioeconomic status (SES; low, medium, high), religion (Jewish or non-Jewish), country of birth (Israel or outside Israel) and periphery residence (southern and northern districts). Body mass index (BMI), smoking status, Charlson Comorbidity Index, and disease history (chronic pulmonary disease, ischemic heart disease, anxiety, depression, opioid use disorder, opioid overdose, heroin overdose, HIV, hepatitis-C, mental health disease, cancer and diabetes). BMI and smoking data were missing for less than 2% of participants, and for those variables, the denominator included patients with non-missing data. History of opioid use was defined as prescription dispensations that occurred before the diagnosis of opioid use disorder. Characteristics were compared using the Wilcoxon signed-rank test for continuous variables and the Chi-square test for discrete variables (p<0.05).

We examined the ratio between the mean healthcare cost of members with opioid use disorder divided by that of the controls, which was translated from the local currency, New Israeli Shekel (NIS), to United States Dollar (at a ratio of 1 NIS = USD 0.31). The costs were averaged over months of exposure. The confidence interval (CI) of this healthcare cost ratio was calculated using the percentile bootstrap method with 10,000 iterations.

As a sensitivity analysis, and in order to focus particularly on populations with prescription opioid use disorder (by excluding heroin use), the ratio was also examined separately among members without HIV, hepatitis C, and heroin poisoning before the index date (as an indicator of probable heroin use). The analysis was also stratified by sex and age (18-64 years vs. 65 years and older). Specific analyses pertaining to different healthcare costs included inpatient costs, emergency room visits, outpatient services, prescription drug purchases, laboratory tests, and imaging. Finally, we analyzed the most common diagnoses for which healthcare services were provided. The analysis was performed using the R statistical package. This study was approved by the CHS institutional review board.

## Results

Opioid dependence was diagnosed at least once in 126 members in 2013, and started to increase after 2015, reaching 271 members by 2019. Opioid abuse was diagnosed in 78 members in 2013, and started to increase after 2016, reaching 168 members by 2019 (Figure 1).

**Figure 1:**
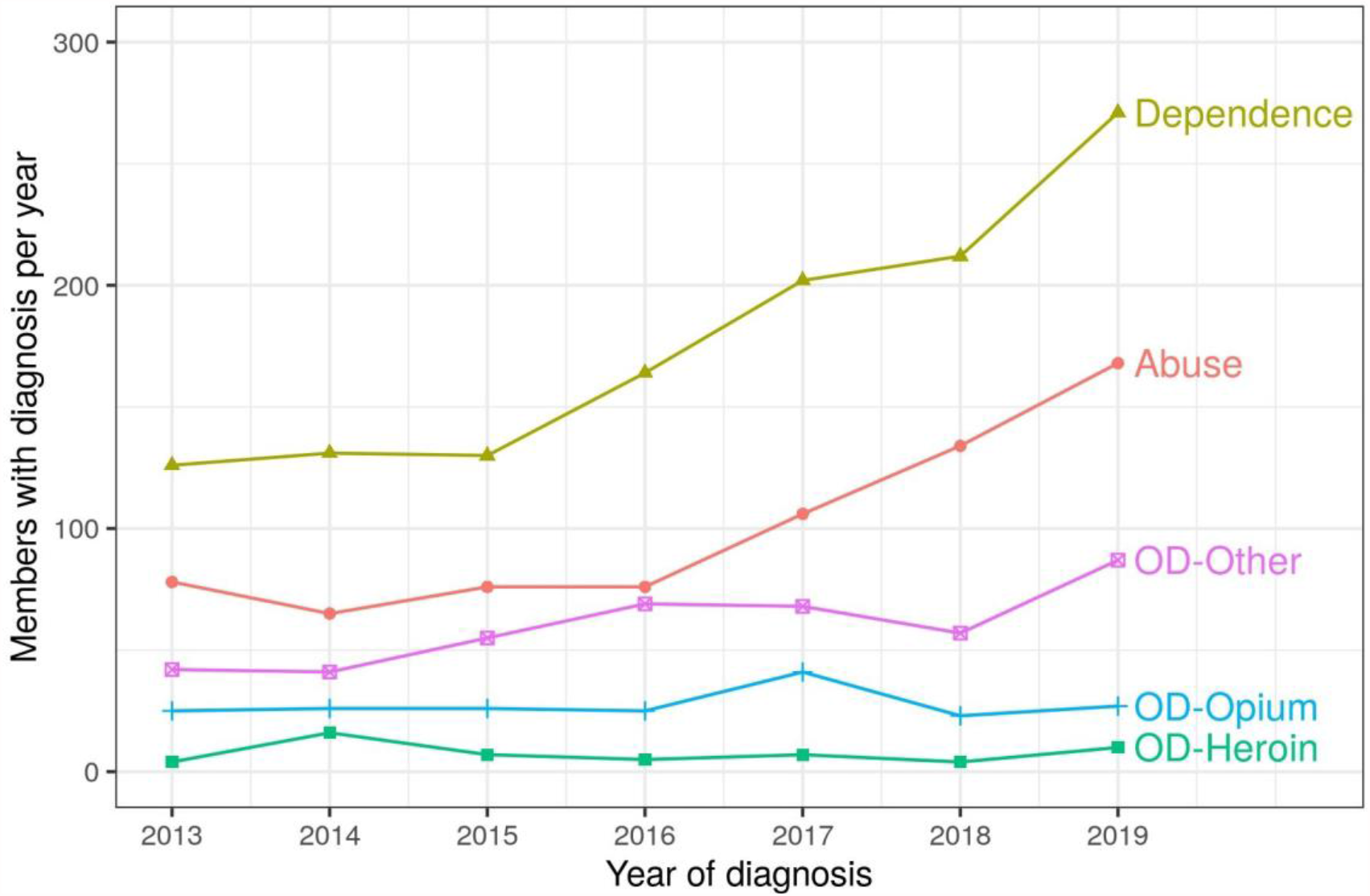
Opioid use diagnoses by year. Legend: The Y axis indicates the number of members with a given diagnosis per year, and the X axis indicates the year of diagnosis. Yellow indicates opioid dependence; red indicates opioid abuse; blue indicates overdose from opium (including morphine); green indicates overdose from heroin; and purple indicates overdose from other opioids (primarily Oxycodone and Fentanyl). Abbreviations: OD, overdose.

Our final study population included 1,173 patients. Patients diagnosed with opioid use disorder were matched to 11,730 controls by sex and age. The majority of the participants were male (74.3%) and had a median age of 51 years (Table 1).

**Table 1:** Demographic characteristics of the patient population.

|  | Opioid use disorder | Controls | P value |
| --- | --- | --- | --- |
| **N** | 1,173 | 11,730 |  |
| **Male (%)** | (74.3) | (74.3) | 1* |
| **Age, median (IQR)** | 51 (39, 60) | 51 (39, 60) | 1* |
| **Age group (%)** |  |  | 1* |
| **18-29** | (7.2) | 7.2 |  |
| **30-39** | 18.2 | 18.2 |  |
| **40-49** | 20.6 | 20.6 |  |
| **50-59** | 27.8 | 27.8 |  |
| **60-69** | 15.4 | 15.4 |  |
| **70-79** | 6.1 | 6.1 |  |
| **≥ 80** | 4.6 | 4.6 |  |
| **Socioeconomic status (%)** |  |  | <0.001 |
| **Low** | 31.8 | 31.6 |  |
| **Medium** | 60.3 | 50.1 |  |
| **High** | 7.9 | 18.3 |  |
| **Jewish (%)** | 88.6 | 84.0 | <0.001 |
| **Immigrant (%)** | 37.7 | 64.4 | <0.001 |
| **Residence in periphery (%)** | 33.8 | 22.0 | <0.001 |

\*Age and sex were used to match the groups
Abbreviations: IQR, interquartile range.

When compared with controls, those with opioid use disorder were approximately 4 times more likely to have a history of mental health disease, to be underweight (BMI below 20 kg/m^2^), and 3 time2ws more likely to smoke. They were also 15 times more likely to have HIV and 30 times more likely to have hepatitis C (Table 2).

**Table 2:** Clinical characteristics of the patient population.

|  | **Opioid use disorder** | **Controls** | **P value** |
| --- | --- | --- | --- |
| **Opioid use disorder history = Yes (%)** | 9.3 | 0.1 | <0.001 |
| **Opioid overdose before index = Yes (%)** | 3.9 | 0 | <0.001 |
| **Heroin overdose before index = Yes (%)** | 1.9 | 0 | <0.001 |
| **Opioid prescription use before index** | 83.0 | 55.9 | <0.001 |
| **Fentanyl prescription use before index** | 21.5 | 0.8 | <0.001 |
| **Mental health disease history = Yes (%)** | 55.2 | 13 | <0.001 |
| **Depression history = Yes (%)** | 37.3 | 8.7 | <0.001 |
| **Anxiety history = Yes (%)** | 43.4 | 13.1 | <0.001 |
| **Body mass index (kg/m<sup>2</sup>), median (IQR)** | 24.2 (21.2, 28.0) | 26.1 (23.4, 29.5) | <0.001 |
| **Smoking = Yes (%)** | 71.3 | 24.1 | <0.001 |
| **Charlson Comorbidity Index (median [IQR])** | 2 [1, 4] | 1 [0, 3] | <0.001 |
| **Chronic pulmonary disease before index = Yes (%)** | 18.5 | 2.8 | <0.001 |
| **Ischemic heart disease before index = Yes (%)** | 13.6 | 9.2 | <0.001 |
| **Cancer before index = Yes (%)** | 11.5 | 5.5 | <0.001 |
| **Diabetes before index = Yes (%)** | 17.2 | 14.7 | 0.022 |
| **Human immunodeficiency virus before index = Yes (%)** | 1.5 | 0.1 | <0.001 |
| **Hepatitis C before index = Yes (%)** | 31 | 0.9 | <0.001 |

Abbreviations: IQR, interquartile range.

The mean monthly healthcare cost for members with opioid use disorder was $1,102 compared to $211 among controls, a 5.2-fold difference (95% CI: 4.6-6.0). After excluding members with HIV, Hepatitis C, or heroin overdose prior to the index date, the monthly healthcare cost of members with opioid use disorder was $979, compared to $124 among controls, a difference of 4.6-fold (95% CI: 3.9-5.4).

For individuals aged 18-64 years (non-elderly adults(, the monthly healthcare cost among those with opioid use disorder was $1049, whereas among controls it was $173, a difference of 6.1-fold (95% CI: 5.2-7.1). For those aged 65 years and older (elderly), the monthly healthcare cost in members with opioid use disorder was $1,369, and in controls it was $399, a difference of 3.4-fold (95% CI 2.6-4.5).

Among men, the monthly healthcare cost in members with opioid use disorder was $1,414, compared to $193 among controls, a difference of 5.6-fold (95% CI: 4.7-6.5). Among women, the monthly healthcare cost for those with opioid use disorder was $1,181 per month, compared to $261 among controls, a difference of 4.5-fold (95% CI: 3.5-5.8).

Among those with high SES, the monthly healthcare cost in members with opioid use disorder was $1,414, which was higher than controls by 5.6-fold (95% CI: 3.7-8.5). Individuals with opioid use disorder in the medium and lower SES groups had lower absolute healthcare costs compared to the high SES group, but had a similar ratio of costs compared to the controls in the same SES group (Figure 2).

**Figure 2:**
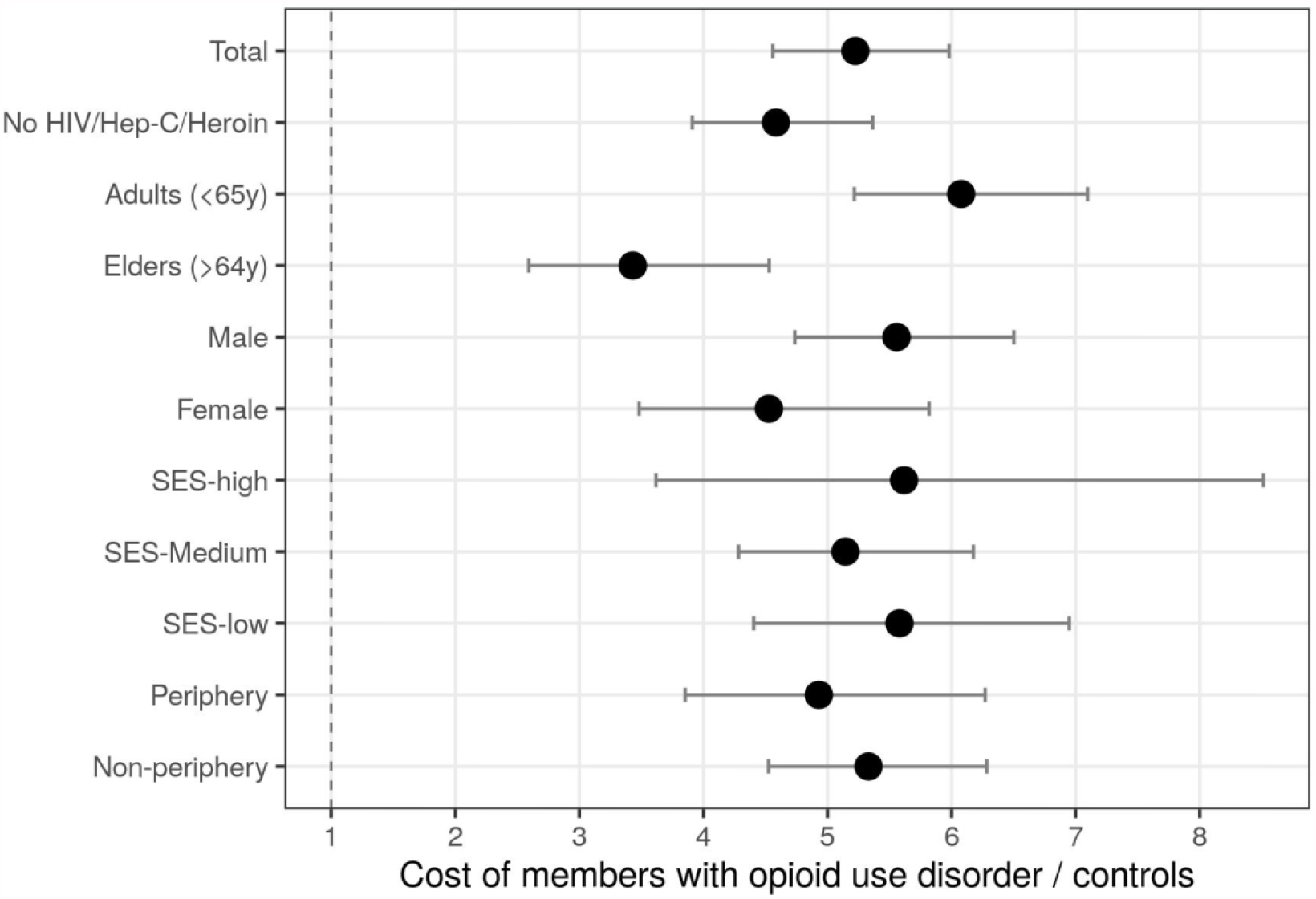
Ratio of healthcare cost among patients with opioid use disorder compared with controls. Legend: The Y axis indicates the inclusion criteria for the analysis and the X axis indicates the healthcare cost of members with opioid use disorder divided by the cost in controls. The circle indicates the ratio and the error bars indicate the 95% confidence intervals. Abbreviations: HIV, Human Immunodefinicy Virus; SES, Socioeconomic Status

The medical service that was associated with the highest cost among those with opioid use disorder was inpatient services, which accounted for an average of $742 a month, and was higher than controls by 8.7-fold (95% CI: 7.2-10.4). The medical service with the second highest cost among those with opioid use disorder was prescription drug dispensing, which accounted for a monthly average of $218, which was higher than controls by 3.3-fold (95% CI: 2.6-4.1; Figure 3).

**Figure 3:**
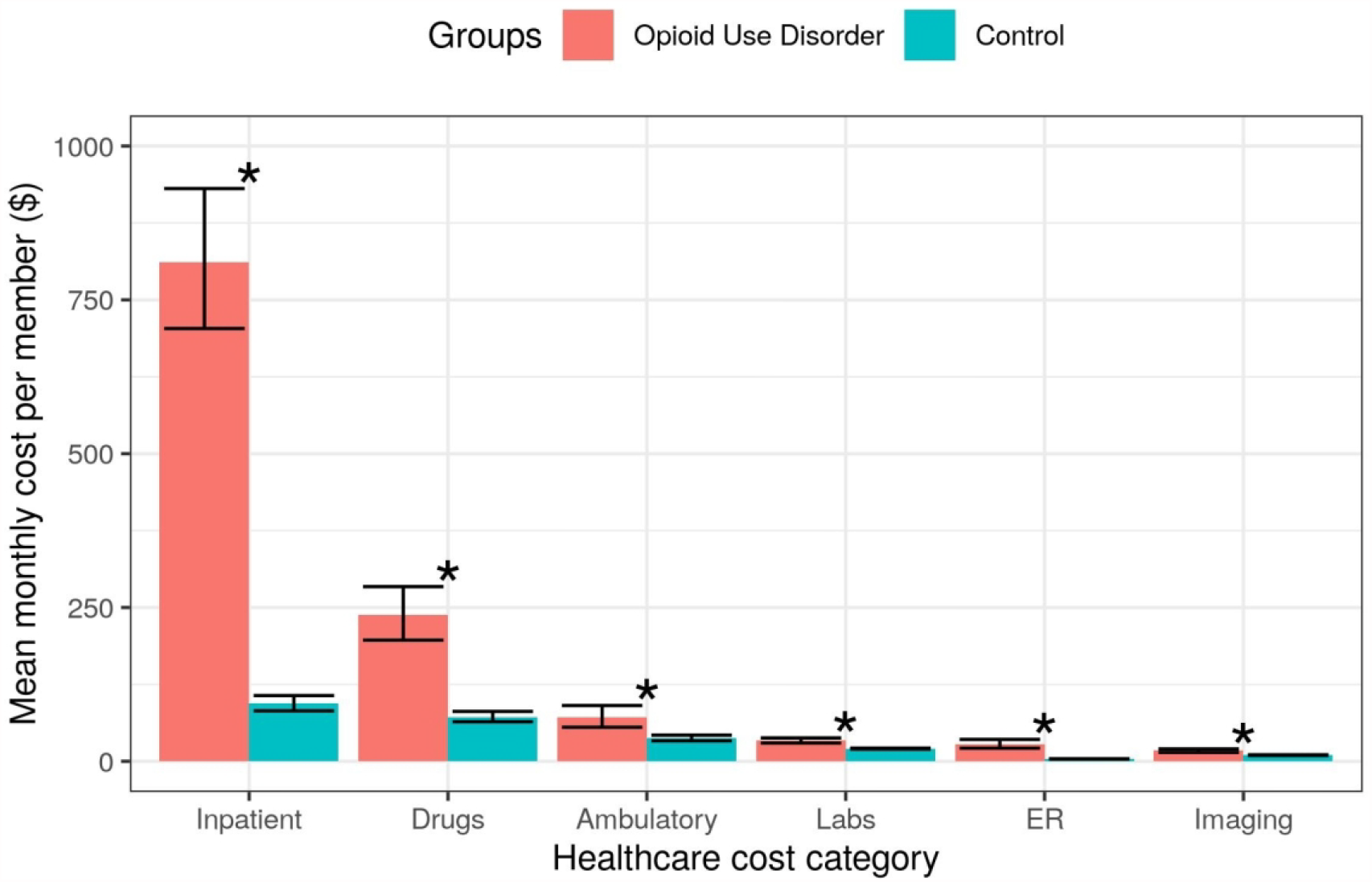
Healthcare cost by service type among members with opioid use disorder compared to controls. Legend: The Y axis indicates mean monthly cost per member and the X axis indicates the healthcare cost category. Red indicates cases with opioid use disorder and blue indicates controls. Error bars indicate the 95% confidence intervals and asterisks denote a category where the cost in the opioid use disorder groups was found to be significantly higher compared to the controls. Abbreviations: ER, emergency room.

After excluding cases with HIV, hepatitis C, or heroin use before the index date, most healthcare costs in members with opioid use disorder were derived from inpatient services ($429/month), which were 7-fold (95% CI: 5.7-8.7) higher than among controls.

Regarding inpatient services, the sub-category with the highest costs was urgent hospitalizations, which accounted for $580 a month among those with opioid use disorder, and was 17.4-fold (95% CI: 14.1-21.6) higher than controls. Elective hospitalizations accounted for $153 a month among those with opioid use disorder, 3-fold (95% CI: 2.1-4.4) higher than among controls.

The most common inpatient diagnosis for members with opioid use disorder included non-dependent abuse of drugs and drug dependence (Figure 4).

**Figure 4:**
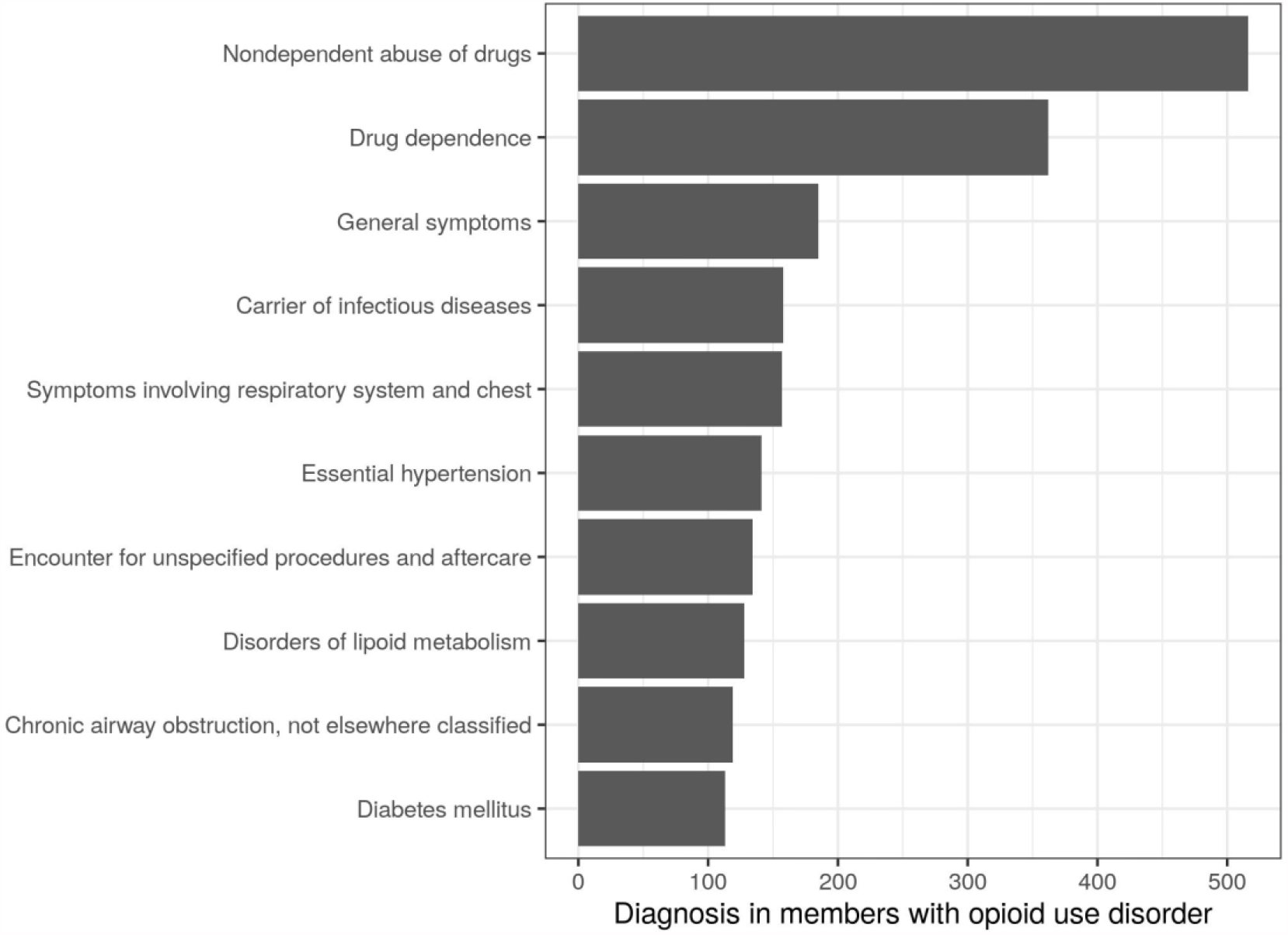
Most common inpatient diagnoses among individuals with opioid use disorder. Legend: The Y axis indicates inpatient diagnosis and the X axis indicates the amount of diagnosis in members with opioid use disorder.

## Discussion

This study compared healthcare costs among individuals with opioid use disorder and their matched controls within a public health system. Our findings indicate a 5-fold difference in healthcare costs associated with opioid use disorder, mostly attributed to inpatient services.

Many of the costs associated with opioid use disorder were attributed to prescription opioids. In line with previous data from the US, the relative healthcare cost difference in members with opioid use disorder was larger among individuals younger than 65 years compared to those 65 or older. This most likely stems from in healthcare costs being higher among older members regardless of concurrent opioid use disorder.[8,13] Previous research has indicated that preventing opioid use disorder at a younger age leads to larger savings in healthcare and societal costs.[7]

The most common healthcare service that was utilized by individuals with opioid use disorder was inpatient services, which was 9 times higher than in controls. The second highest service among those with opioid use disorder was emergency room services (7-fold), which has also been shown as a major source of healthcare costs among those with opioid use disorders in the US.[14] The most common inpatient and emergency room diagnosis for those with opioid use disorders was drug abuse. Another common diagnosis was respiratory symptoms, which could relate in part to the respiratory depression that can be caused by opioids.[15] Costs related to inpatient services have been less often cited in connection with opioid use disorder than opioid prescriptions. Health services and policy makers should account for these differences in costs when considering investments in prevention and treatment of opioid use disorder.

A potential limitation of our study concerns the proportion of members with a diagnosis code for opioid use disorder: we expected a higher number of individuals to be diagnosed with opioid use disorder, based on previous studies that have estimated its prevalence in Israel and globally.[16,17] The relatively low rate is also unexpected considering recent police reports of widespread opioid abuse in Israel, and the recent finding of increased opioid consumption within CHS.[10,18]. Similar difficulties in identifying opioid use disorder and probable under-reporting has been shown in other countries as well.[19] The probable under-diagnosis of opioid use disorder in the Israeli healthcare services records could be attributed to treatment being under the direct clinical and financial care of the Israeli Ministry of Health and not through the healthcare organizations. Another possible factor for the low rates is the relatively low awareness regarding prescription opioid abuse among physicians in Israel. Nevertheless, although the rates of opioid use disorder were most likely under-reported in this study, the comparison of healthcare costs was made possible by comparing those with a positive diagnosis as “true positive”, and those without it as “true negative” (the matched controls). Since most of the population does not have opioid use disorder, and our sample used 10 controls for each case, the effect of false negatives is unlikely to have a drastic influence on our findings.

Another limitation of the study is that the root illness that was driving the initial use of opioids may in some cases be associated, by itself, with long-term increased healthcare utilization that cannot be easily separated from the increase attributable to the opioid use. This confounding limits our ability to infer a causal association between opioid use and added healthcare utilization costs.

To conclude, our findings suggest that public healthcare systems incur substantial costs as a result of opioid use disorders, findings which are in line with previous results shown in privatized healthcare systems, such as in the US. It is prudent that public healthcare systems improve their identification of members with opioid use disorder, by raising awareness among clinicians and including systemic alerts to decrease unwarranted prescription of these medications. Early intervention and referral to specialized treatment should be implemented in public healthcare systems. In addition to clear health benefits, such efforts to better prevent, identify, and treat opioid use disorder could contribute to substantially lowering extensive healthcare costs in this population.

## Supporting information

STROBE checklist

## Data Availability

Owing to data privacy regulations, the raw data for this study cannot be shared.

## Contributors

OM, NB, RB, and SL designed the study. Data were acquired and analyzed by OM. NB, RB, and SL contributed to the analysis. OM wrote the manuscript, and NB, RB, and SL evaluated and edited the manuscript. All authors contributed to and approved the final report. SL is the study guarantor.

## Ethical approval

The study was approved by Institutional Review Board of Clalit Health Services and was exempt from requiring informed consent.

The manuscript’s guarantor (SL) affirms that the manuscript is an honest, accurate, and transparent account of the evaluation being reported; that no important aspects of the evaluation have been omitted; and that any discrepancies from the evaluation as originally planned (and, if relevant, registered) have been explained.

## References

1 McCabe SE, West BT, Veliz P, et al. Trends in Medical and Nonmedical Use of Prescription Opioids Among US Adolescents: 1976-2015. Pediatrics 2017;139. doi:10.1542/peds.2016-2387

2 Rudd RA, Aleshire N, Zibbell JE, et al. Increases in Drug and Opioid Overdose Deaths-United States, 2000-2014. Am J Transplant 2016;16:1323–1327. doi:10.1111/ajt.13776

3 Dahan A, Aarts L, Smith TW. Incidence, Reversal, and Prevention of Opioid-induced Respiratory Depression. Anesthesiology 2010;112:226–238. doi:10.1097/ALN.0b013e3181c38c25

4 Des Jarlais DC, Arasteh K, Feelemyer J, et al. Hepatitis C virus prevalence and estimated incidence among new injectors during the opioid epidemic in New York City, 2000-2017: Protective effects of non-injecting drug use. Drug Alcohol Depend 2018;192:74–79. doi:10.1016/j.drugalcdep.2018.07.034

5 McCarty D, Perrin NA, Green CA, et al. Methadone maintenance and the cost and utilization of health care among individuals dependent on opioids in a commercial health plan. Drug Alcohol Depend 2010;111:235–240. doi:10.1016/j.drugalcdep.2010.04.018

6 Mohlman MK, Tanzman B, Finison K, et al. Impact of Medication-Assisted Treatment for Opioid Addiction on Medicaid Expenditures and Health Services Utilization Rates in Vermont. J Subst Abuse Treat 2016;67:9–14. doi:10.1016/j.jsat.2016.05.002

7 Murphy SM. The cost of opioid use disorder and the value of aversion. Drug Alcohol Depend 2020;217:108382. doi:10.1016/j.drugalcdep.2020.108382

8 White AG, Birnbaum HG, Mareva MN, et al. Direct costs of opioid abuse in an insured population in the United States. J Manag Care Pharm 2005;11:469–479. doi:10.18553/jmcp.2005.11.6.469

9 Ponizovsky AM, Marom E, Weizman A, et al. Changes in consumption of opioid analgesics in Israel 2009 to 2016: An update focusing on oxycodone and fentanyl formulations. Pharmacoepidemiol Drug Saf 2018;27:535–540. doi:10.1002/pds.4415

10 Miron O, Zeltzer D, Shir T, et al. Rising opioid prescription fulfillment among non-cancer and non-elderly patients—Israel’s alarming example. Regional Anesthesia & Pain Medicine 2020.

11 Chang H-Y, Kharrazi H, Bodycombe D, et al. Healthcare costs and utilization associated with high-risk prescription opioid use: a retrospective cohort study. BMC Med 2018;16:69. doi:10.1186/s12916-018-1058-y

12 Shei A, Hirst M, Kirson NY, et al. Estimating the health care burden of prescription opioid abuse in five European countries. Clinicoecon Outcomes Res 2015;7:477–488. doi:10.2147/CEOR.S85213

13 Pasquale MK, Joshi AV, Dufour R, et al. Cost drivers of prescription opioid abuse in commercial and Medicare populations. Pain Pract 2014;14:E116–25. doi:10.1111/papr.12147

14 Langabeer JR, Stotts AL, Bobrow BJ, et al. Prevalence and charges of opioid-related visits to U.S. emergency departments. Drug Alcohol Depend 2021;221:108568. doi:10.1016/j.drugalcdep.2021.108568

15 Dahan A, Aarts L, Smith TW. Incidence, reversal, and prevention of opioid-induced respiratory depression. The Journal of the American Society of … 2010.

16 Degenhardt L, Grebely J, Stone J, et al. Global patterns of opioid use and dependence: harms to populations, interventions, and future action. Lancet 2019;394:1560–1579. doi:10.1016/S0140-6736(19)32229-9

17 Nigmalim. Estimate:’ ‘ 15,000 Israelis are addicted to pain killers. 2017.

18 Tara Kavaler. Everyone Knows Somebody“: The opioid epidemic among Israel”s orthodox. Jerusalem Post. 2019.

19 Lagisetty P, Garpestad C, Larkin A, et al. Identifying individuals with opioid use disorder: Validity of International Classification of Diseases diagnostic codes for opioid use, dependence and abuse. Drug Alcohol Depend 2021;221:108583. doi:10.1016/j.drugalcdep.2021.108583

